# Assessing Predictors of Mortality Among Children admitted with Sepsis at a Referral Tertiary Health Center, Northwestern Nigeria

**DOI:** 10.1101/2022.08.04.22278417

**Authors:** Fatimah Hassan-Hanga, Baffa Sule Ibrahim, Halima Kabir, U Hafsat Ibrahim, Kabiru Abdulsalam, Zainab Datti Ahmed, Halima Salisu Kabara, Sule Abdullahi Gaya, Dalha Gwarzo Haliru, Nasiru Magaji Sadiq, Salisu Inuwa, Mohammad Aminu Mohammad

**Author notes:** **Corresponding author:** Baffa Sule Ibrahim.

## Abstract

**Background:** Sepsis is a life-threatening infection that can lead to organ failure and death. We aim to assess predictors of mortality among children admitted with Septicemia at a referral health facility in Northwestern Nigeria.

**Methods:** We conducted a prospective cross-sectional study of children aged 0-14 years admitted to various units of the pediatrics department of the health facilities. Children were recruited between September 2018 and November 2019. All recruited children were followed up on clinical progress until either discharge, abscondment, or death. We assessed the children clinically daily and collected whole-blood samples for laboratory tests. We conducted a univariate and multivariable analysis using STATA-16 to assess identified predictive factors with our outcome variable.

**Results:** A total of 326 children were recruited, median age: 2-years. About 54.0% of the children were boys, and 53.1% were within 1-5 years age-group. Predominant organisms cultured from the blood of the children were *Salmonella typhi* (5.7%), *Klebsiella pneumoniae* (2.3%), and *Staphylococcus aureus* (2.0%). A total of 35 deaths were recorded with a case fatality rate (CFR) of 10.7%. CFR is highest in children <1years (13.6%).

Child’s vaccination status, mother’s education level as well as blood lactate levels, GCS, qSOFA score and positive blood culture were significantly associated with child’s mortality. Factors associated with increase mortality include; children with incomplete vaccination history [OR=1.72, 95%CI: 2.74–15.53] versus those with full vaccination; children whose mothers had no formal education [OR=14.39, 95%CI: 3.24–63.99] when compared to those children whose mothers have tertiary level of education. Furthermore, children with whole blood lactate level between 4-8mmol/l [OR=3.23, 95%CI: 1.15–9.07], or greater than 8mmol/l [OR=10.54, 95%CI: 3.68–30.14] versus children with whole blood lactate level less than 4mmol/l; children with qSOFA score of 3 [OR=15.62, 95%CI: 3.31–73.60] versus children with qSOFA score of 1; and children who had a positive blood culture [OR=6.90, 95%CI: 3.04–15.64].

**Conclusion:** We found a high prevalence of severe sepsis at pediatrics department of AKTH. Serum lactate levels, GCS, and qSOFA scores were predictive of mortality. Routine measurement and monitoring of these parameters will improve case management and reduce sepsis related mortality in the hospital.

## Introduction

Sepsis is one of the main causes of death and morbidity worldwide, with a particularly devastating impact in low-income nations. Tissue damage, organ failure, and death are all possible outcomes of the body’s excessive and life-threatening response to infection.^[1][2][3]^ Pediatric sepsis is a term that refers to a group of illnesses caused by bacteria, viruses, fungi, or parasites, as well as the harmful products of these pathogens. Infants and children benefit greatly from early detection and intervention^[4][5]^. Sepsis-causing infections are a significant cause of death and morbidity in children worldwide^[6]^, It affects people of all ages and is caused by a variety of infectious diseases, including pneumonia, malaria, meningitis, tuberculosis (TB), HIV, and COVID-19^[2,7]^.

Sepsis affects all age groups, accounting for 20% of global deaths^[8]^. In 2017, children accounted for half of all sepsis cases worldwide, with an estimated 20 million cases and 2.9 million fatalities in under 5yrs children. An estimated 17 million of these cases and 3.5 million of the deaths occurring in Africa^[9][10]^. Sepsis also accounts for 15% of neonatal deaths, and is the most common cause of death in infants globally. According to the World Health Organization, sepsis due to severe pneumonia, severe diarrhea, severe malaria, and severe measles, are responsible for the highest deaths in children^[11]^.

Significant regional disparities in Sepsis incidence and mortality exist between high income (HIC) and low-and middle-income countries (LMIC)^[7,8,12]^, with 85% of sepsis cases and related deaths occurring in Africa^[13]^. Studies have shown that, children in LMICs are 18 times more likely to die under the age of five than children in higher income countries (HICs)^[7,8,12]^. The observed discrepant survival may be caused by the interplay between socio-economic, factors and poor medical care and resource limitations^[6]^. Nigeria is the second highest contributor to under-five mortality in the world, majority of these deaths were due to sepsis^[14]^.

Several factors are reported to be associated with severe sepsis in children. In the United States, infants are at highest risk of sepsis, with a rate ten times higher than that of older children^[15]^. About 25% of severe sepsis occurs in Low-and very low-birth-weight (VLBW) children. Also, boys less than 10yrs of age had significantly higher rates of severe sepsis than girls, particularly among infants^[15]^.

Blood culture remains the gold standard in the microbiological diagnosis of sepsis, however, other prognostic scoring systems, laboratory investigations and biomarkers play a key role^[16]^. Early identification and understanding the signs and symptoms of sepsis, along with the detection of important prognostic biomarkers (such as blood lactate and procalcitonin), are crucial elements for early diagnosis of sepsis and the timely establishment of its appropriate clinical management. Illness scoring systems (such as the Glasgo Coma Score, and the Quick Sequential Organ Assessment Score) are widely used in critically ill patients^[17]^. However, their use in children with sepsis has largely been limited and shredded in ambiguity^[18]^.

Biomarkers can be classified based on their clinical application. Prognostic biomarkers provide information on the likely outcome in an untreated child and help in establishing the intensity of treatment. Monitoring biomarkers serve to evaluate the disease process and are especially informative of the effectiveness of response to the therapeutic intervention, allowing the clinician to titrate or change the management protocol^[16,19]^.

Early intervention of sepsis especially in children is critical, and delays in commencing treatment worsen prognosis. Even HICs, with the most advanced healthcare systems struggle to deliver recommended care in sepsis timely^[20]^. With Nigeria facing similar challenges to other African countries with poor infrastructure and delivery system, its important for clinicians to have an established protocol for early detection of prognostic parameters in pediatrics sepsis. This study aimed to review and assess prognostic factors for mortality in admitted children at a referral tertiary health center in North western Nigeria, and to evidently advocate for a review of the current sepsis management protocol.

## Methods

### Study Design

We conducted a prospective cross-sectional study on children (ages 0-14 years) who were admitted with sepsis to pediatrics unit of Aminu Kano Teaching Hospital (AKTH), from the first of September 2018 to 30th November 2019. Enrolled children were subsequently evaluated clinically until they were either discharge, absconded or dead.

### Sampling

All admitted children were eligible. Enrolment of consenting patients for the study was done for all eligible children. Enrollment commenced form the 1^st^ of September and continued until four weeks prior to end of the study.

### Study Population

Children ages 0-14 years admitted with sepsis to pediatrics department of AKTH Kano.

### Data Collection

A pre-tested interviewer administered questionnaire was used to collect patient information from the parents or guardians and entered into Open Data Kit (ODK) using smart phones. The following socio-demographic information were collected; hospital ID, name, age, sex, date of admission, ward of admission, parents’ occupation and educational levels, child’s vaccination history, and ethnicity. A team of experts (Pediatricians) conducted clinical examination on the enrolled children and documented signs and symptoms of sepsis. Clinical parameters recorded include Glasgow Coma Score (GCS), and the Quick Sequential Organ Failure Assessment (qSOFA).

Anthropometric parameters (weight, height, mid upper-arm circumference, head circumference) and vital signs (body temp, respiratory rate, heart rate) were measured and recorded at enrollment. At lease 4ml of whole blood was collected from peripheral vein of each child after disinfection using iodine solution, then left to dry and wiped off with (70%) alcohol. Blood culture was done using automated BACTEC^®^ 9050 blood-culture system (Becton Dickinson, Temse, Belgium) for a maximum of 5 days. About 2mls of the blood samples were inoculated into a commercially produced vials containing Fastidious Antimicrobial Neutralization Plus media (FAN^®^ Plus) for aerobic and anaerobic media, and incubated in the BACTEC^®^ incubator. The FAN^®^ Plus media contains resins (Adsorbent Polymeric Beads) which absorb out prior antibiotics used, thus allowing the growth of organisms and enhancing the recovery of isolates. Positive vials were then aliquoted and sub-cultured onto Blood, Chocolate and MacConkey agar plates. Isolated pathogens were gram-stained and results recorded. The remaining 2mls were sent (in tubes containing EDTA) to both Microbiology and Chemical Pathology departments for analysis. Experienced laboratory scientists tested for malaria using RDT kit, thick and thin film microscopy. Complete blood count was done using automated 3-parts hematology analyzer. Estimation for whole blood lactate were done using Roche Accutrend plus^®^ point of care testing (POCT) analyzer. Urea, Electrolytes and Creatinine were determined using the Abbott Architect c4000^®^ chemistry analyzer.

### Data Management and Storage

Patients’ information were collected using pre-tested questionnaires via open data kit software pre-installed into secured smartphones. Data were backed up daily to a secured laptop stored (in csv format) at a different location. Data entry was monitored daily and corrected for errors, and a final data cleaning was done at the end of the study.

### Data Analysis

Statistical analysis was done using STATA-16. Descriptive statistics were used to summarize frequencies and percentages for categorical data and median and interquartile range for continuous data. In bivariate analysis, comparison of proportions was performed using Chi-square and exact Fisher’s test, P-value of less than 0.05 was considered as statistically significant. Logistic regression was used to analyze sociodemographic and sepsis risk factors associated with clinical outcome. We conducted a manual backward stepwise regression to identify significant factors for the regression models. Two regression models were done. A reduced model (only included factors identified as significant using the backwards stepwise regression) and the full model which combined all socio-demographic, clinical and laboratory factors, adjusting for potential confounders and eliminating multicollinearity.

### Ethical Clearance

The study protocol and all data collection tools were approved by Research Ethical Review Sub-committee of the Kano State Ministry of Health, with approval number: MOH/Off/797/T.I/732, written and dated 1^st^ June 2018.

## Results

### Socio-demographic characteristics

We recruited a total of 326 eligible children between September 2018 and November 2019. The median age was 2 years, with age range of 0 – 14 years. There were 176 (54.0%) boys and 150 (46.0%) girls. About 53.1% of the children were within the 1 – 5 years age bracket, while 27.0% were under one year. Majority, 296 (90.8%) of the patients were enrolled from the Emergency Pediatrics Unit (EPU) while 3 (0.9%) and 9 (2.8%) were enrolled from Oncology and Pediatrics Medical Wards (PMW) respectively. Also, most of the patients 284 (89.3%) were Hausa, while Yoruba and Igbo tribes constituted 7 (2.8%) and 8 (3.2%) respectively. Among recruited children, 165 (53.4%) were fully vaccinated, 90 (29.1%) had incomplete vaccination, while 54 (17.5%) had no any history of vaccination. Thirty-five (35) deaths were recorded (**Table 1**).

**Table 1:**
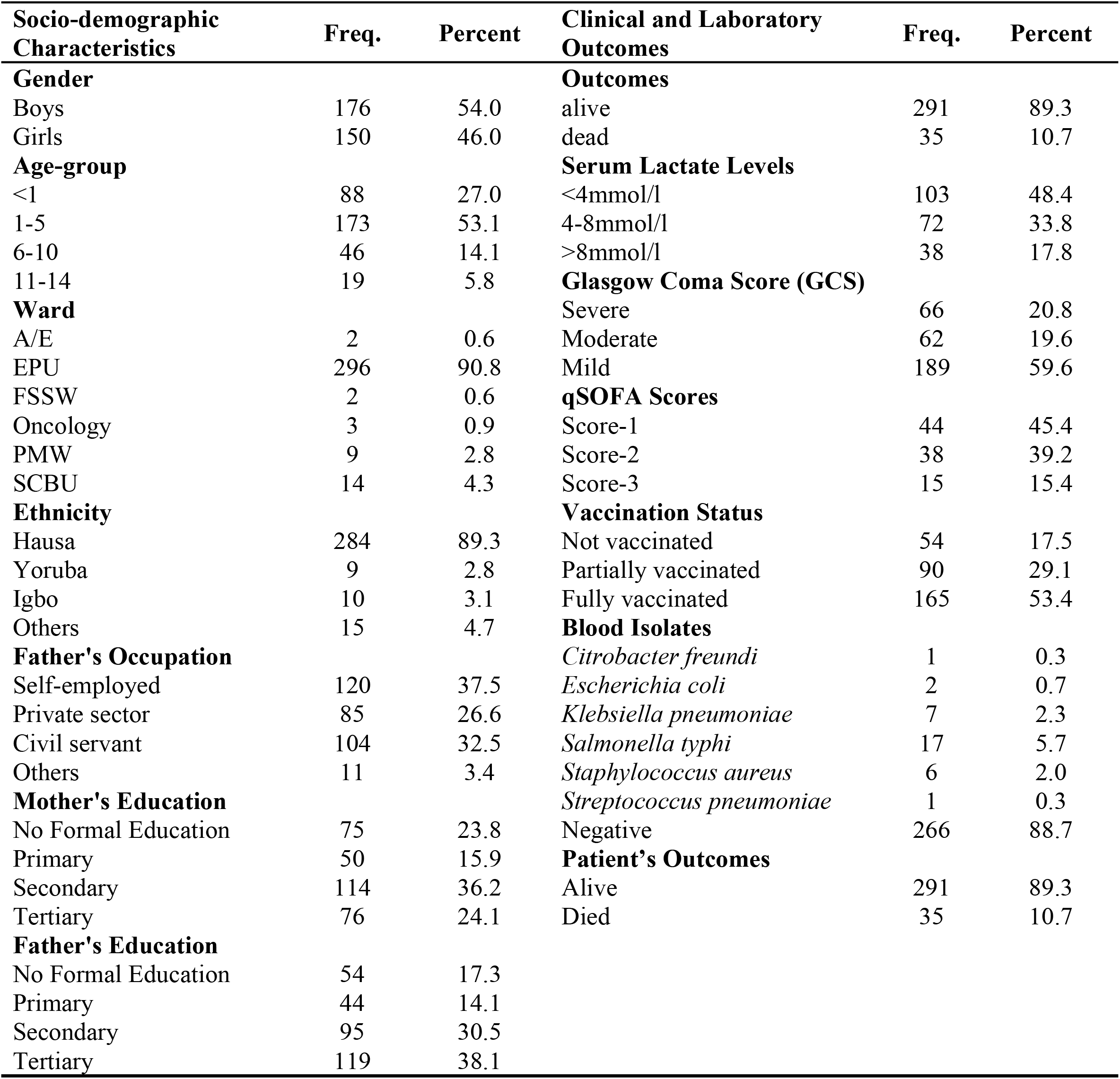
Distribution of Socio-demographic characteristics, clinical and laboratory outcomes of children (aged 0 – 14years) admitted with sepsis at pediatrics units of AKTH, Kano (n=326).

| **Socio-demographic Characteristics** | **Freq.** | **Percent** | **Clinical and Laboratory Outcomes** | **Freq.** | **Percent** |
| --- | --- | --- | --- | --- | --- |
| **Gender** |  |  | **Outcomes** |  |  |
| Boys | 176 | 54.0 | alive | 291 | 89.3 |
| Girls | 150 | 46.0 | dead | 35 | 10.7 |
| **Age-group** |  |  | **Serum Lactate Levels** |  |  |
| <1 | 88 | 27.0 | <4mmol/l | 103 | 48.4 |
| 1-5 | 173 | 53.1 | 4-8mmol/l | 72 | 33.8 |
| 6-10 | 46 | 14.1 | >8mmol/l | 38 | 17.8 |
| 11-14 | 19 | 5.8 | **Glasgow Coma Score (GCS)** |  |  |
| **Ward** |  |  | Severe | 66 | 20.8 |
| A/E | 2 | 0.6 | Moderate | 62 | 19.6 |
| EPU | 296 | 90.8 | Mild | 189 | 59.6 |
| FSSW | 2 | 0.6 | **qSOFA Scores** |  |  |
| Oncology | 3 | 0.9 | Score-1 | 44 | 45.4 |
| PMW | 9 | 2.8 | Score-2 | 38 | 39.2 |
| SCBU | 14 | 4.3 | Score-3 | 15 | 15.4 |
| **Ethnicity** |  |  | **Vaccination Status** |  |  |
| Hausa | 284 | 89.3 | Not vaccinated | 54 | 17.5 |
| Yoruba | 9 | 2.8 | Partially vaccinated | 90 | 29.1 |
| Igbo | 10 | 3.1 | Fully vaccinated | 165 | 53.4 |
| Others | 15 | 4.7 | **Blood Isolates** |  |  |
| **Father's Occupation** |  |  | *Citrobacter freundii* | 1 | 0.3 |
| Self-employed | 120 | 37.5 | *Escherichia coli* | 2 | 0.7 |
| Private sector | 85 | 26.6 | *Klebsiella pneumoniae* | 7 | 2.3 |
| Civil servant | 104 | 32.5 | *Salmonella typhi* | 17 | 5.7 |
| Others | 11 | 3.4 | *Staphylococcus aureus* | 6 | 2.0 |
| **Mother's Education** |  |  | *Streptococcus pneumoniae* | 1 | 0.3 |
| No Formal Education | 75 | 23.8 | Negative | 266 | 88.7 |
| Primary | 50 | 15.9 | **Patient's Outcomes** |  |  |
| Secondary | 114 | 36.2 | Alive | 291 | 89.3 |
| Tertiary | 76 | 24.1 | Died | 35 | 10.7 |
| **Father's Education** |  |  |  |  |  |
| No Formal Education | 54 | 17.3 |  |  |  |
| Primary | 44 | 14.1 |  |  |  |
| Secondary | 95 | 30.5 |  |  |  |
| Tertiary | 119 | 38.1 |  |  |  |

### Clinical and laboratory findings

Out of the 326 enrolled children, 103 (48.4%) had whole blood lactate levels of <4.0mmol/l, while 72 (33.8%) and 38 (17.8%) had whole blood lactate levels between 4-8mmol/l and >8mmol/l respectively. Also, Glasgow Coma Score (GCS) at enrollment was mild in 59.6%, moderate in 19.6%, and severe in 20.8% of the children. Quick Sequential Organ Failure Assessment (qSOFA) revealed a score-1 in 45.4%, score-2 in 39.2% and score-3 in 15.4% of the children that had the assessment. (**Table 1**).

Severe malaria (103), pneumonia (54) and meningitis (48) constituted the most prevalent causes of sepsis at the center (**Figure 1**). *Salmonella typhi* (5.7%), *Klebsiella pneumoniae* (2.3%), and *Staphylococcus aureus* (2.0%), were the predominant organisms cultured from the blood of the children over the study period (**Table 1**).

**Figure 1.**
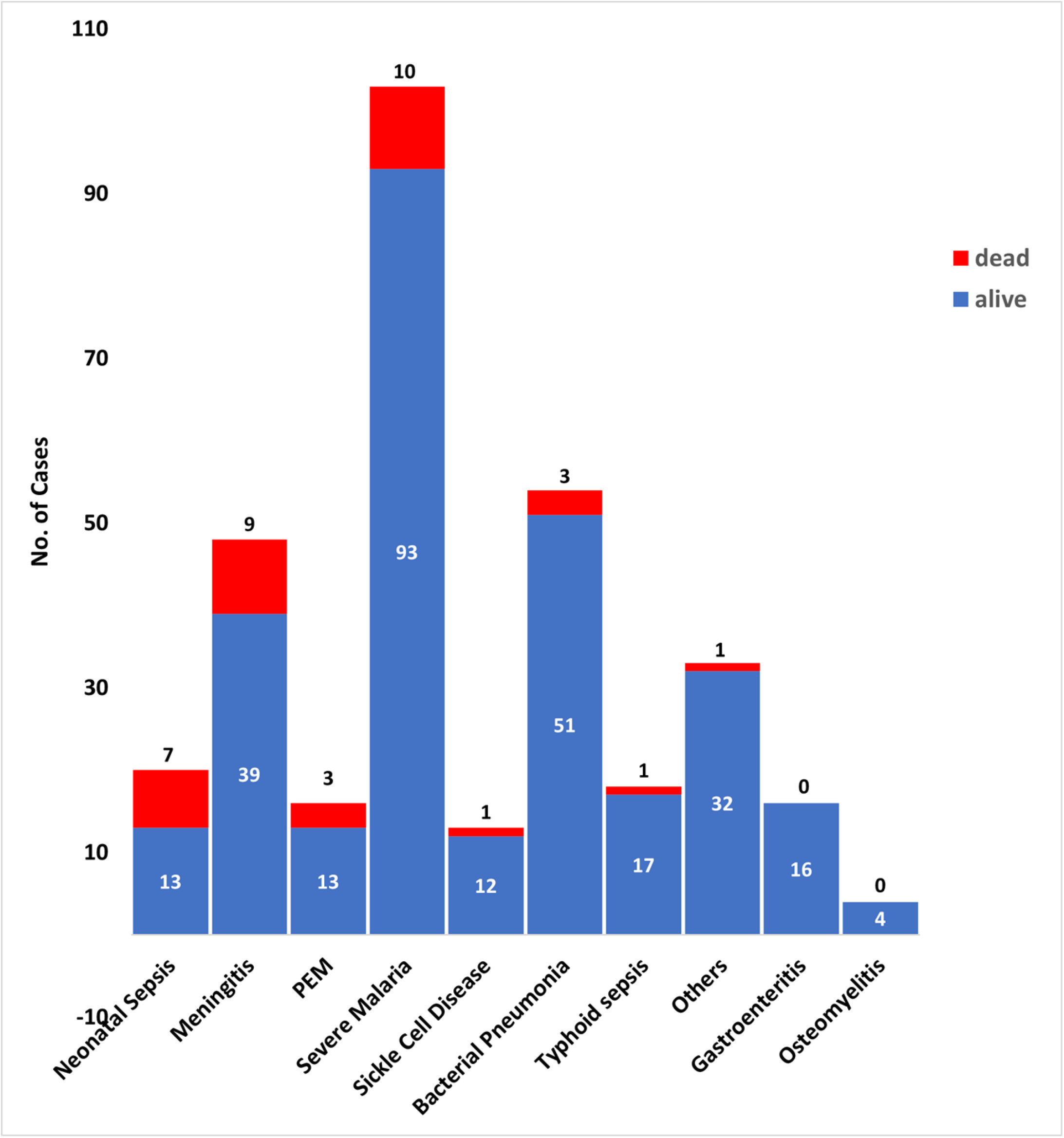
Clinical diagnosis disaggregated by outcome of children (aged 0 – 14years) admitted with sepsis at pediatrics units of AKTH, Kano (n=326).

Of the 326 enrolled children, 35 deaths were recorded which gives a case fatality rate (CFR) for sepsis of 10.7%. Disease specific fatality rate (DSFR) was highest in children diagnosed with neonatal sepsis (35.0%), followed by those diagnosed with Cerebrospinal meningitis (18.8%) and PEM (18.8%). There were no recorded deaths in children diagnosed with sepsis due to gastroenteritis or osteomyelitis (**Table 2**).

**Table 2:** Distribution of disease specific fatality rate (DSFR) and age specific fatality rate (ASFR) among children (aged 0 – 14years) admitted with sepsis at pediatrics units of AKTH, Kano (n=326)

| **Clinical diagnosis** | **Patient's outcomes** |  |  | **DSFR** |
| --- | --- | --- | --- | --- |
|  | **alive** | **dead** | **Total** |  |
| Neonatal Sepsis | 13 | 7 | 20 | 35.0% |
| Meningitis | 39 | 9 | 48 | 18.8% |
| PEM | 13 | 3 | 16 | 18.8% |
| Severe Malaria | 93 | 10 | 103 | 9.7% |
| Sickle Cell Disease | 12 | 1 | 13 | 7.7% |
| Bacterial Pneumonia | 51 | 3 | 54 | 5.6% |
| Typhoid sepsis | 17 | 1 | 18 | 5.6% |
| Others | 32 | 1 | 33 | 3.0% |
| Gastroenteritis | 16 | 0 | 16 | 0.0% |
| Osteomyelitis | 4 | 0 | 4 | 0.0% |

| **Age-groups** | **Patient's outcomes** |  |  | **ASFR** |
| --- | --- | --- | --- | --- |
|  | **alive** | **died** | **Total** |  |
| <1yr | 76 | 12 | 88 | 13.6% |
| 1-5yrs | 154 | 19 | 173 | 11.0% |
| 6-10yrs | 43 | 3 | 46 | 6.5% |
| 11-14yrs | 18 | 1 | 19 | 5.3% |

### Bivariate and Multivariate

In bivariate analysis, the following factors were significantly associated with high chance of mortality among the children: child’s vaccination status (p<0.001), mother’s education (p<0.001), and father’s education (p<0.001) (**Table 3**). Likewise, whole blood lactate levels (p<0.001), GCS (p<0.001), qSOFA score (p<0.001) and blood culture (p<0.001) were found to be significantly associated with increase mortality among the enrolled children (**Table 4**).

**Table 3:**
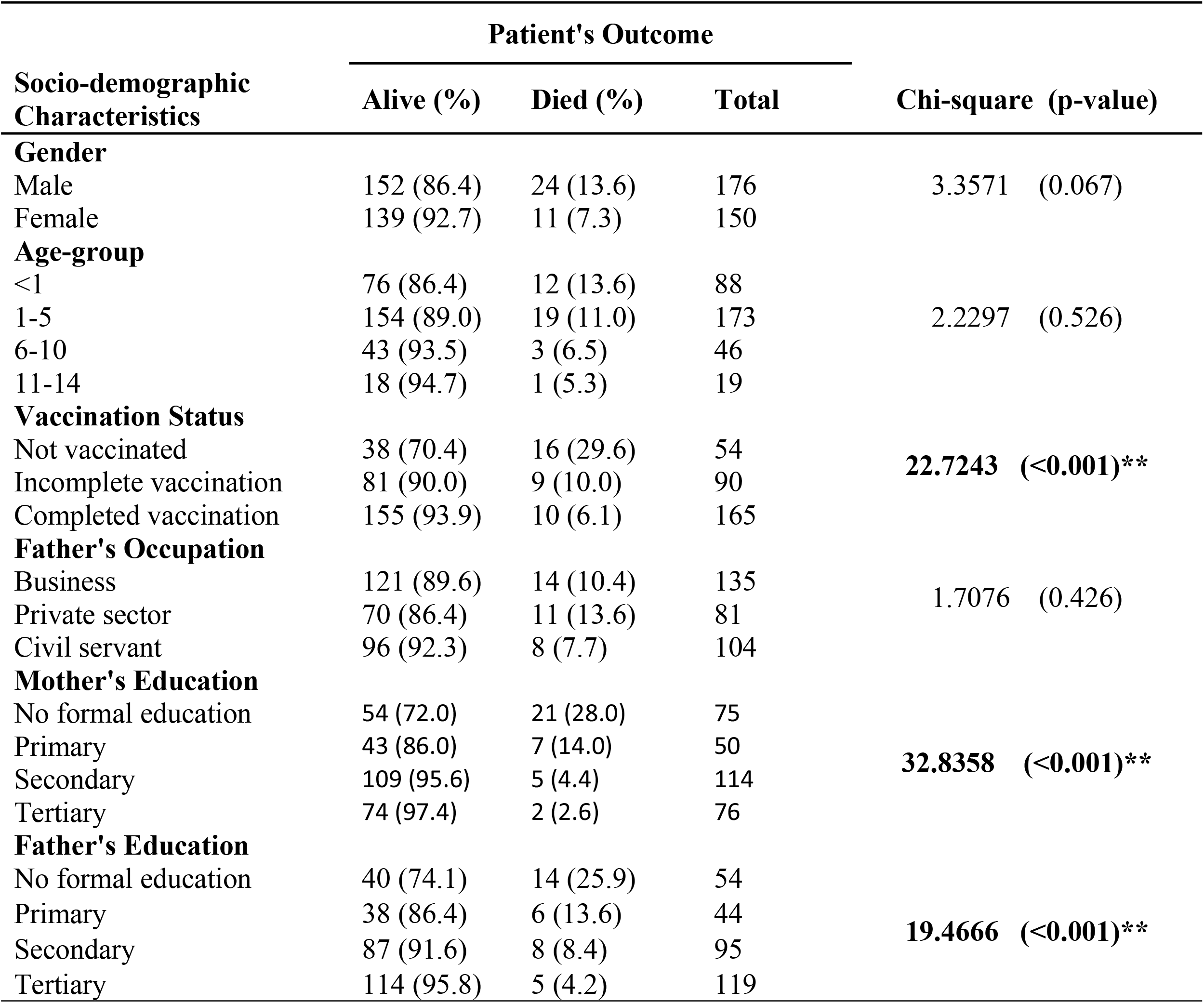
Bivariate relationships between socio-demographic characteristics and patient’s outcome among children (aged 0 – 14years) admitted with sepsis at pediatrics units of AKTH, Kano (n=326).

**Table 4:** Bivariate relationships between clinical and laboratory findings and patient’s outcome among children (aged 0 – 14years) admitted with sepsis at pediatrics units of AKTH, Kano (n=326).

| Clinical and Laboratory Outcomes | Patient's Outcome |  |  | Chi-square (p-value) |
| --- | --- | --- | --- | --- |
|  | Alive (%) | Died (%) | Total |  |
| Serum Lactate Levels |  |  |  |  |
| <4mmol/l | 97 (94.2) | 6 (5.8) | 103 | 24.1196 (<0.001)** |
| 4-8mmol/l | 60 (83.3) | 12 (16.7) | 72 |  |
| >8mmol/l | 23 (60.5) | 15 (39.5) | 38 |  |
| Glasgow Coma Scale (GCS) |  |  |  |  |
| Severe | 51 (77.3) | 15 (22.7) | 66 | 16.6885 (<0.001)** |
| Moderate | 53 (85.5) | 9 (14.5) | 62 |  |
| Mild | 179 (94.7) | 10 (5.3) | 189 |  |
| qSOFA Scores |  |  |  |  |
| Score-1 | 41 93.2) | 3 (6.8) | 44 | 27.0768 (<0.001)** |
| Score-2 | 33 (86.8) | 5 (13.2) | 38 |  |
| Score-3 | 7 (46.7) | 8 (53.3) | 15 |  |
| Duration on admission |  |  |  |  |
| <=24hrs | 28 (93.3) | 2 (6.7) | 30 | 6.7076 (0.082) |
| 2-6days | 64 (75.3) | 21 (24.7) | 85 |  |
| 7-14days | 11 (64.7) | 6 (35.3) | 17 |  |
| >14days | 6 (66.7) | 3 (33.3) | 9 |  |
| Blood Culture Result |  |  |  |  |
| Positive for blood pathogen | 23 (63.9) | 13 (36.1) | 36 | 26.3493 (<0.001)** |
| Negative for blood pathogen | 244 (92.4) | 20 (7.6) | 264 |  |

**Table 5** and **Table 6** contain results of the full and reduced regression models for the socio-demographic and clinical outcomes respectively. In the reduced model for socio-demographic, several factors were associated with increase mortality among the patients: children with incomplete vaccination history [OR=1.72, 95% CI: 2.74–15.53] versus those with full vaccination; children whose mothers have no formal education [OR=14.39, 95% CI: 3.24–63.99] when compared to those children whose mothers have tertiary level of education; children whose fathers have no formal education [OR=7.89, 95% CI: 2.70–23.56] when compared to those children whose fathers have tertiary level of education.

**Table 5:**
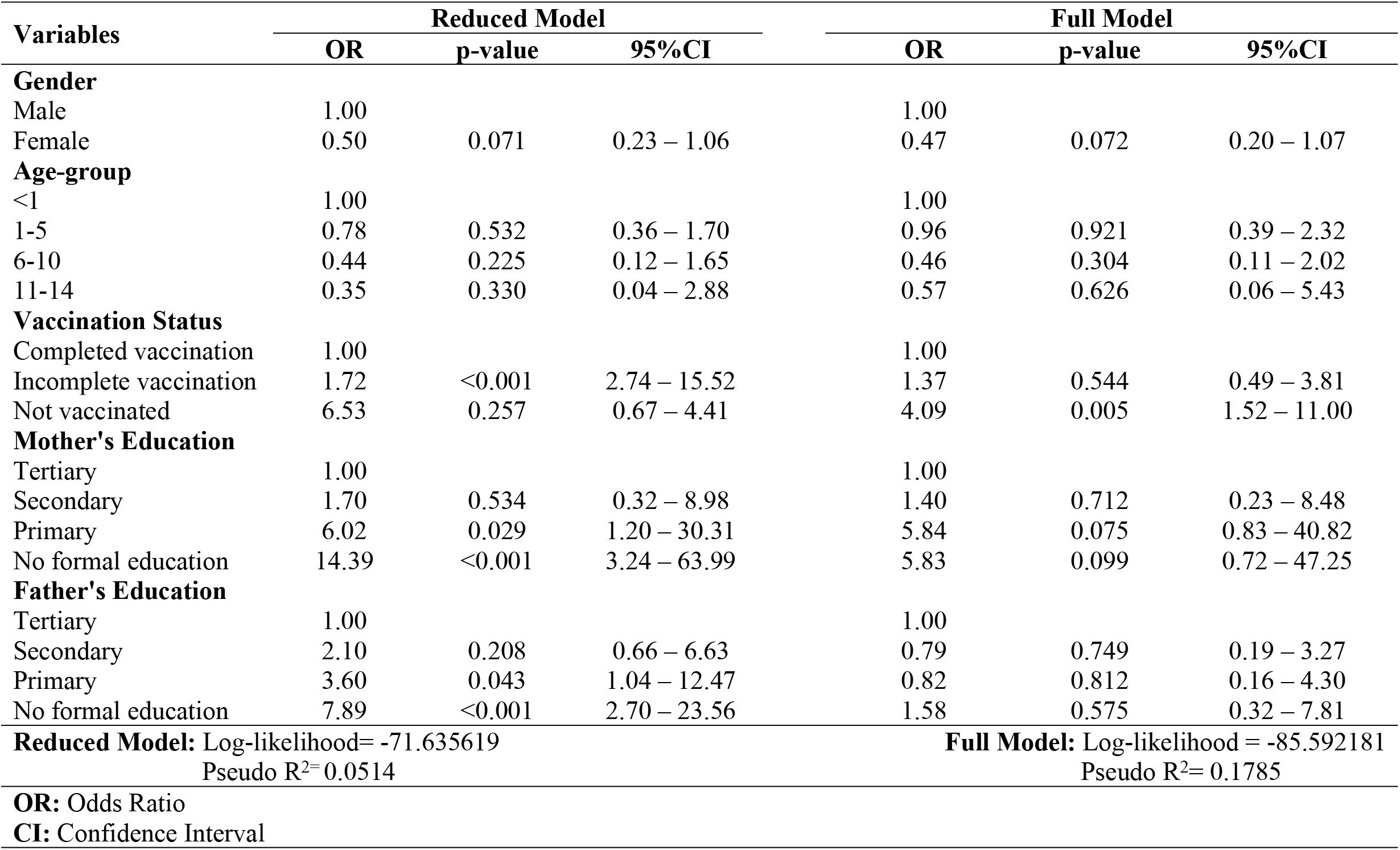
Multivariate logistic regression models with odds rations, P-value and confidence intervals for socio-demographic factors, for children (aged 0 – 14years) admitted with sepsis at pediatrics units of AKTH, Kano (n=326).

**Table 6:**
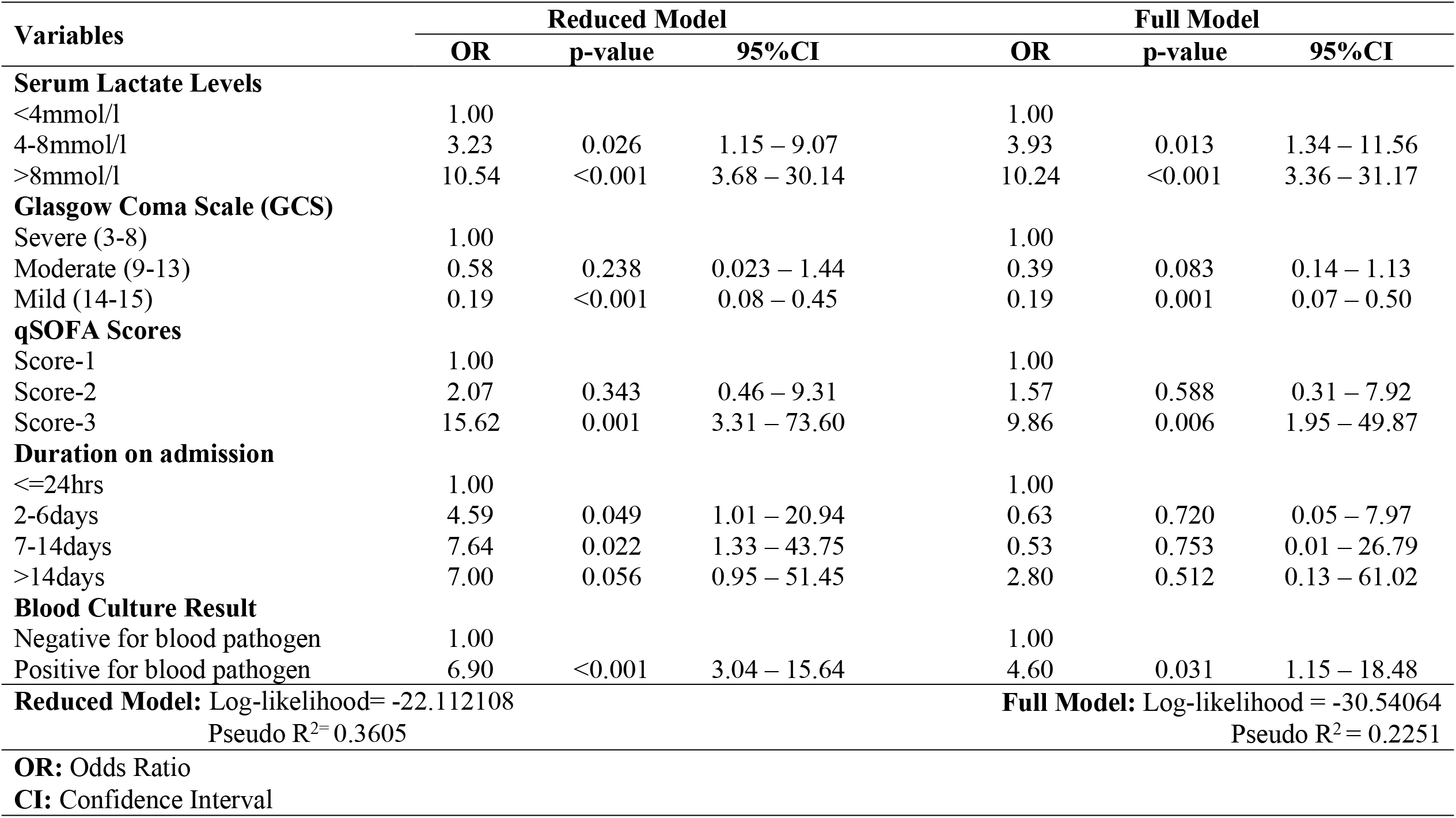
Multivariate logistic regression models with odds rations, P-value and confidence intervals for clinical and laboratory outcomes, for children (aged 0 – 14years) admitted with sepsis at pediatrics units of AKTH, Kano (n=326).

In the reduced model for the clinical and laboratory outcomes, several factors were also associated with increase mortality among the patients: children with whole blood lactate level between 4-8mmol/l [OR=3.23, 95% CI: 1.15–9.07], or greater than 8mmol/l [OR=10.54, 95% CI: 3.68– 30.14] versus children with whole blood lactate level less than 4mmol/l; children with qSOFA score of 3 [OR=15.62, 95% CI: 3.31–73.60] versus children with qSOFA score of 1; children who spend between 2-6days on admission [OR=4.59, 95% CI: 1.01–20.94]], or 7-4days on admission [OR=7.64, 95% CI: 1.33–43.75], compared to children who spend less than 24hrs on admission; children who have a positive blood culture [OR=6.90, 95% CI: 3.04–15.64]. A GCS score between 14-15 (mild) at enrollment is associated with improved survival [OR=0.19, 95% CI: 0.08–0.45] when compared to children with GCS of 3-8 (severe) at enrolment.

## Discussion

This study found case fatality rate for sepsis to be 10.7%. This was comparable to similar studies in Nigeria^[21,22]^ and studies by El-Mashad et al in Egypt^[23]^ where the mortality rate was 8.8% but lower than 22% reported by Novosad et al^[24]^ and 23.8% reported in Saudi Arabia^[25]^. This difference could be due to difference in the studied population where their patients were those admitted into the pediatric intensive care unit only, indicating the level of severity of the disease. Majority of our patients were males (59.9%) and within 1-5 years of age (53.3%). A quarter (25.9%) of the patients were less than one year of age. This is similar to many Nigerian studies where most of their patients were less than five years^[21,22,26]^. It is also similar to the study by Novosad et al where 39% of their patients were less than one year^[24]^. This was expected as sepsis has a biphasic peak, first in early childhood and the second peak in the elderly.

Only 58.1% of our patients were fully immunized, which is less than the 67% reported in the 2018 Nigerian National Nutrition and Health Survey^[27]^. This is worrisome because many of the sepsis cases are from infectious diseases. In this study, malaria (30.7%), Pneumonia (16.7%) and Meningitis (14.8%) were the most prevalent causes of sepsis. This was similar to a study conducted in Saudi Arabia^[25]^ where the respiratory system was the most common infection site but different from Novosad et al^[24]^ where most of their patients had congenital heart disease and cerebral palsy. This also reflects the difference in epidemiology between patients in high income countries where chronic diseases are more prevalent and those from low-and middle-income countries (LMIC) where infectious diseases are still more prevalent. Vaccination has been proven to be a highly effective prevention strategy against many infectious diseases in children like Pneumonia, meningitis, Tuberculosis etc.

Neonatal sepsis had the highest disease fatality rate in our study, this was similar to findings in Abakaliki (23.9%)^[28]^ and Jos (27.3%)^[29]^ but lower than that obtained in Ile-Ife (33.3%)^[30]^. This finding also strengthens the fact that sepsis is one of the commonest causes of hospitalization among neonates. Meningitis and severe acute malnutrition followed next with disease fatality rates of 11.8% and 11.1% respectively. This was similar to findings by Ling et al where they identified malnutrition as one of the risk factors for sepsis^[31]^.

There was significant association between mortality and not being immunized, having high serum lactate, poor Glasgow Coma Score (GCS) and a high qSOFA score. Blood lactate levels has been used to measure disease severity in patients with sepsis/septic shock. This finding was similar to a study by Hatherill et al that showed that hyperlactemia and its persistence after 24 hours of treatment can predict death^[32]^. Though high Serum lactate levels have been associated with increased mortality especially in adult patients presenting with septic shock, some studies (23,14) in children have not been so conclusive. Gorgis et al did not find any association between an elevated early lactate and mortality in children with sepsis but it however correlated strongly with PRISM-III, a measure of mortality risk in pediatrics^[33]^. In this study, poor GCS was significantly associated with mortality, this was similar to findings by Crippa et al who carried out a post hoc analysis of intensive care over Nations database investigating the effect of brain dysfunction, measured by the GCS on hospital mortality in critically ill patients^[34]^. They concluded that hospital mortality was significantly higher in patients with brain dysfunction measured with GCS and particularly in the presence of sepsis^[34]^.

Higher qSOFA scores were found to be associated with increased mortality in this study, similar to the study by EL-Mashad et al where they found that the score was higher in non-survivors and the mortality increased progressively across their patient subgroups from lower to higher scores^[23]^. However, some studies have reported that the qSOFA score has a poor sensitivity, 37%-70% in predicting in-hospital mortality^[35,36]^.

In our study, *Salmonella typhi, Staphylococcus aureus* and *Klebsiella pneumonia* were the predominant organisms cultured, this was similar to findings by Obaro et al where Salmonella species account for 24%-59.8% of bacteremia in Central and Northwest Nigeria. This however, differs from findings by Ogunkunle et al where *Staphylococcus aureus* was the most predominant organism isolated from their patients^[21]^. This further signifies that there is a significant variation in the bacterial organisms causing sepsis even within the same region and country.

## Conclusion

Pediatric sepsis is common in our hospital with a case fatality rate of 10.7%. Mortality from sepsis was significantly associated with not being fully immunized, poor GCS and high serum lactate levels. Neonates were more affected while in older children, infectious diseases like malaria, Pneumonia and meningitis were the predominant causes. *Salmonella typhi, Staphylococcus aureus* and *Klebsiella spp* were the predominant organisms cultured.

## Data Availability

All datasets are available and will be provided to request to lead author

## Data Availability

Dataset for this study is available on request to the corresponding author (address above) or the PI of the study: Dr. Fatima Hasan-Hanga, +234 803 714 1639.

## Acknowledgements

The authors will like to acknowledge and appreciate the Kano State Ministry of Health, AKTH management, all clinical and nursing departments, and our team of data entrants for their unfiltered support and dedication to the completion of this study.

## Authors Contributions

Dr. Fatima Hassan-Hanga contributed to the conception and design of the study and acquisition of data. She also contributed to revising the manuscript for important intellectual content and final approval of the version to be published.

Dr. Baffa Sule Ibrahim contributed to conception and design of the study, acquisition of data, data analysis and interpretation. He substantially contributed to drafting the article and final approval of the version of the manuscript to be published.

Dr. Halima Kabir, Dr. Hafsat Ibrahim, Dr. Kabiru Abdulsalam, Dr. Zainab Datti Ahmed, Halima Salisu Kabara, Sule Abdullahi Gaya, Dalha Gwarzo Halliru, Nasiru Magaji Sadiq, Salisu Inuwa, and Prof. Mohammad Aminu Mohammad, all contributed to the conception and design of the study, acquisition of data, revising the manuscript for important intellectual content, and approval of the version to be published.

